# Overuse in US Medicare during the COVID-19 pandemic: 2020 versus 2019

**DOI:** 10.1101/2022.05.25.22275006

**Authors:** Kelsey Chalmers, Shannon Brownlee, Valérie Gopinath, Vikas Saini

**Author notes:** Corresponding author: Kelsey Chalmers.

## Abstract

**Background:** The COVID-19 pandemic and March 2020 shutdown in the US reduced the volume of healthcare services, but the impact on overuse has not been investigated.

**Objective:** To examine the change in overuse rates and volumes through 2020.

**Design:** A retrospective cohort study using Medicare fee-for-service claims.

**Setting:** Outpatient and inpatient claims.

**Participants:** Patients who met the criteria for one of 10 overuse measures with a claim between January 1 2019 to December 31 2020.

**Measurements:** Overuse volumes were reported as patients with claims meeting overuse metric criteria per 100,000 Medicare beneficiaries. Overuse rates were measured by the same overuse cohort per 100 patients meeting the denominator criteria of the metric. Rates in 2020 were compared to the same date period in 2019 using incidence rate ratios (IRRs) estimated from Poisson regressions.

**Results:** In 2019, 302,379 patients had an overuse claim (14.72% of 2,053,792 patients in the cohort) versus 234,481 (13.79% of 1,699,807) in 2020. The overall cohort included 2,112,904 (61.0%) women and a mean (SD) age of 76.5 (8.1) years. There was a 52.3% decrease in overall cohort volume during the COVID-19 shutdown; 2,341,017 patients in 2020 versus 4,912,453 in 2019. There was a 72.57% decrease in patients with an overuse procedure between April 2019 (N = 11,794) and 2020 (N = 3,220) (IRR 0.27 (95% CI 0.25 to 0.3; p <0.001)), including spinal fusion/laminectomy, carotid endarterectomy, knee arthroscopy, hysterectomy and vertebroplasty.

**Limitations:** This study uses claim-based measures of overuse and is limited to the first ten months of the COVID-19 pandemic.

**Conclusions:** The shutdown period during March through May in 2020 had a drastic impact on both the overuse volume and rates for these 10 overuse metrics.

## Introduction

The COVID-19 pandemic caused drastic disruption to usual medical care around the world. During 2020 in the United States, hospitals were inundated with COVID-19 patients from April to June and this triggered a national stay at home order. Infection among hospital personnel meant staff shortages and many hospitals had temporary bed shortages.^1^ While devastating on many levels, the resulting scarcity of resources during this time may have reduced the use of overused or low-value services. Overuse involves the use of tests or procedures that risk patient harm beyond the potential patient benefits and increase health care spending without improving health outcomes.^2^ Estimates of health care overuse have been published on data prior to the COVID-19 pandemic and have found substantial variation in its use across US regions.^3,4^ In this article, we investigated geographic and time variation in overuse in the US traditional Medicare population during 2020.

Previous research has shown that there was a substantial decrease in surgical procedures and hospital admissions in the US during 2020.^5,6^ This decrease occurred even in counties where there were low levels of COVID-19 cases relative to other parts of the country.^6^ Surgery volumes vastly decreased compared to 2019 levels during the initial shutdown period from March 2020.^5^ We therefore expected to find a decrease in overall patient and service volume.

We investigated whether health care providers and patients avoided overuse more than or the same amount as other care during the COVID-19 pandemic. We used a set of 10 overuse measures^7^ and examined changes in the rates of overuse and in the volumes of underlying care delivered.

## Methods

This retrospective cohort study used nationwide traditional Medicare fee-for-service claims to examine rates and frequencies of services during the 2020 COVID-19 pandemic year compared with 2019. The WCG Institutional Review Board approved this study. This study followed Strengthening the Reporting of Observational Studies in Epidemiology (STROBE) reporting guideline for cohort studies.

### Data sources

We used a 100% sample of Medicare fee for service inpatient and outpatient claims from January 1, 2019 to December 31, 2020. We excluded claims if the beneficiary was younger than 65 at the date of service or did not live in the US, or if claims were from facilities not in the 50 US states or D.C.

We used Centers for Disease Control and Prevention (CDC) weekly COVID-19 case data^8^ to distinguish COVID-19 surge periods in states. We defined the initial COVID-19 shutdown period as March 15 to May 2, 2020. This was the same period selected by Mattingly, et al^5^, which encompassed the period where most states had directives to postpone elective surgical procedures. After this period, states were defined as high COVID-19 incidence if the seven-day COVID-19 incidence rate was greater than 175 cases per 100,000 people in the state, and a low COVID-19 incidence otherwise. Like Smulowitz, et al^6^, we selected the threshold of 175 cases per 100,000 population based on the CDC’s hot spot standard of seven-day incidence.

### Measures

We included 10 overuse measures, which we provide in supplementary table 1. The overuse measures we used have two components: a denominator (defined by procedure or diagnosis codes) and a numerator (the subset of the denominator cohort meeting the overuse criteria). For example, the denominator cohort for low-value carotid imaging for syncope included all patients with claims where syncope was recorded as a primary diagnosis on the claim and no exclusion diagnosis codes were recorded (supplementary table 1). The overuse numerator for this measure was any patient within this cohort with carotid imaging.

We used overuse measures for the following services: hysterectomy, knee arthroscopy, carotid endarterectomy, coronary stenting, spinal fusion/laminectomy, vertebroplasty; or a claim for a person presenting with syncope or headache (with some exclusions, the full criteria list is in supplementary table 1).

We measured the volume of overuse as the number of patients with an overuse claim per Medicare FFS beneficiary population, and the rate of overuse as the number of patients with an overuse claim per number in the overuse measure denominator.

### Statistical analysis

We calculated incidence rate ratios (IRRs) with 95% confidence intervals using Poisson regressions. For each measure, we found the total state beneficiary counts who met the measure cohort definition during each period (month or state COVID-19 period) in 2020 as well as the corresponding period in 2019. In the regressions, we included an offset of the log of the state counts of the number of enrolled fee-for-service Medicare beneficiaries who were 65 or older. We then found the state beneficiary counts during the same periods who met the overuse criteria and included in these regressions an offset of the definition denominator. The regression standard errors were clustered at the week and state level.

Statistical significance was assessed at the level of P<.05, and P-values were 2-sided. Statistical analysis was performed using SAS Enterprise Guide version 7.15 and visualizations in R statistical software version 4.0.5 (R Project for Statistical Computing). Data were analyzed from January to March 2022, and project code is available at github.com/Lown-Institute/paper-covid-lvc.

## Results

The study cohort included 3,464,994 beneficiaries in one of the 10 denominator cohorts: 2,053,792 in 2019 and 1,699,807 in 2020. There were 2,112,904 (61.0%) women and the mean (SD) age was 76.5 (8.1) years.

Table 1 gives the cohort counts by year. The overall volume of claims included in denominators decreased by 18.9% in 2020 from 2019, and the overuse volume decreased by 23.4%. Northeastern states had the largest decrease in denominator cohort volumes and overuse volumes compared to other states, while Western states had the smallest decrease in overuse volumes.

**Table 1:** Cohort characteristics and volume comparisons between 2019 and 2020.

| Characteristic | Claims meeting denominator criteria |  |  | Claims meeting low-value service criteria |  |  |
| --- | --- | --- | --- | --- | --- | --- |
|  | 2019 | 2020 | Change (%) | 2019 | 2020 | Change (%) |
| All claims, n | 3,968,716 | 3,218,475 | -18.9 | 339,307 | 259,928 | -23.4 |
| Sex, n (%) |  |  |  |  |  |  |
| Female | 2,401,661<br>(61) | 1,936,783<br>(60) | -19.4 | 191,690<br>(56) | 144,835<br>(56) | -24.4 |
| Male | 1,567,055<br>(39) | 1,281,692<br>(40) | -18.2 | 147,617<br>(44) | 115,093<br>(44) | -22.0 |
| Age, n (%) |  |  |  |  |  |  |
| 65 to 74 | 1,816,421<br>(46) | 1,503,549<br>(47) | -17.2 | 152,458<br>(45) | 118,805<br>(46) | -22.1 |
| 75 to 84 | 1,406,780<br>(35) | 1,132,952<br>(35) | -19.5 | 127,834<br>(38) | 97,640<br>(38) | -23.6 |
| 85 and over | 745,515 (19) | 581,974 (18) | -21.9 | 59,015<br>(17) | 43,483<br>(17) | -26.3 |
| Census region, n (%) |  |  |  |  |  |  |
| Midwest | 984,246 (25) | 826,109 (26) | -16.1 | 78,327<br>(23) | 61,609<br>(24) | -21.3 |
| Northeast | 695,737 (18) | 552,724 (17) | -20.6 | 55,527<br>(16) | 40,510<br>(16) | -27.0 |
| South | 1,531,076<br>(39) | 1,219,265<br>(38) | -20.4 | 150,845<br>(44) | 114,738<br>(44) | -23.9 |
| West | 757,657 (19) | 620,377 (19) | -18.1 | 54,608<br>(16) | 43,071<br>(17) | -21.1 |

### Procedure volumes and overuse rates

Figure 1 shows the 2020 versus 2019 IRRs for each service. The largest changes were in April, and Table 2 presents the counts and rates for this month. There was a 72.57% decrease in the number of patients in April 2020, during the shutdown period, who received a low-value procedure and a 36.01% decrease in the overuse rates per measure denominators (Table 2).

**Figure 1.**
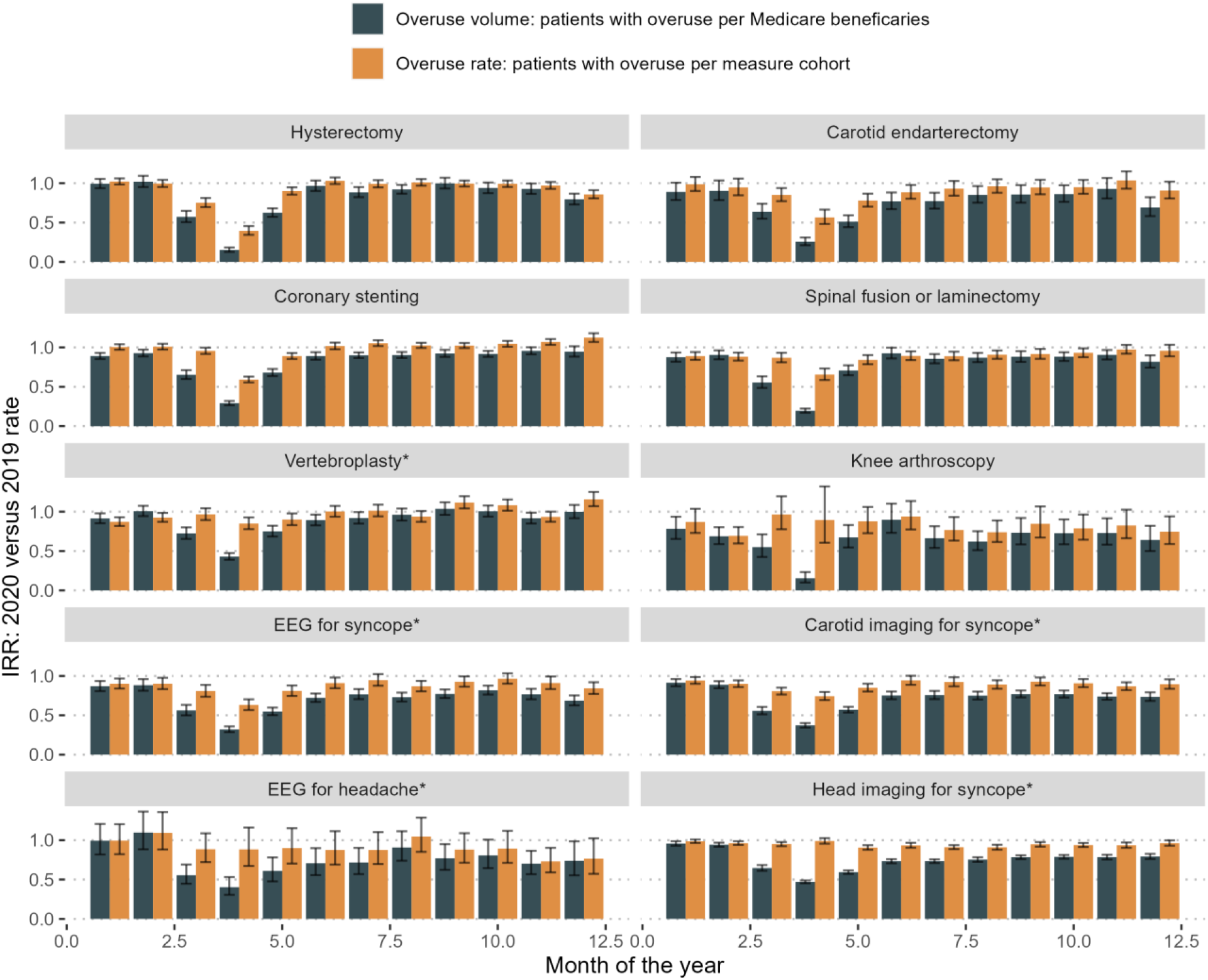
Monthly 2020 versus 2019 incidence rate ratios for overuse measures. The monthly incidence rate ratios (IRRs) for the number of patients with an overuse service per Medicare Fee-For-Service beneficiary (left panel) and per measure denominator cohort (right panel) in 2020 compared to 2019. *Measures where the cohort is defined as claims for patients with a certain condition are marked with an asterisk (vertebroplasty, EEG for syncope, carotid imaging for syncope, head imaging for syncope and EEG for headache patients). Other measures define the cohort as all patients with the specific service. IRRs and 95% CIs (error bars) were estimated from Poisson regressions by comparing total beneficiary counts during epidemiologic weeks in 2020 with corresponding weeks in 2019.

**Table 2:**
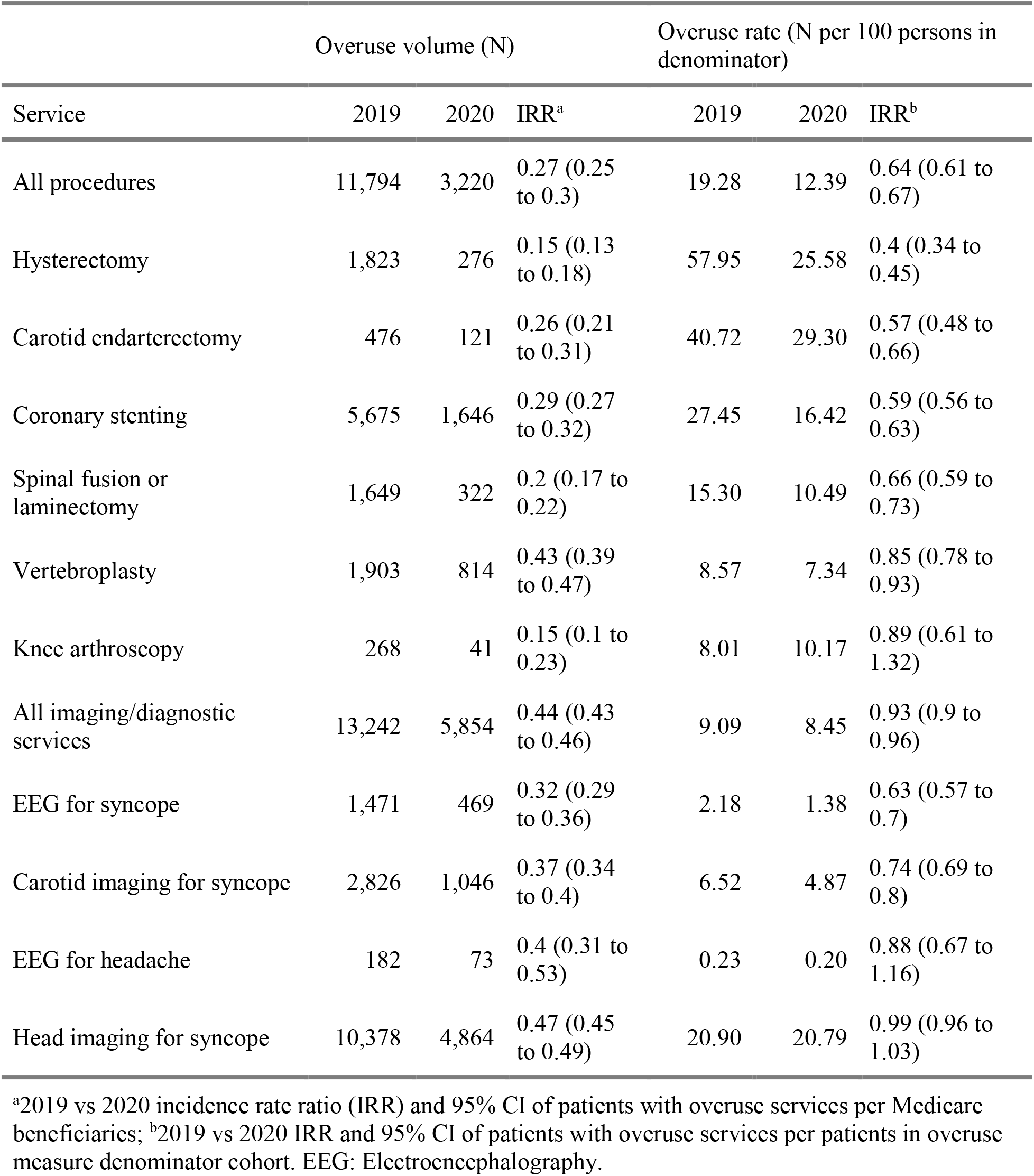
Service overuse counts and rates for April 2019 versus 2020.

| Service | Overuse volume (N) |  |  | Overuse rate (N per 100 persons in denominator) |  |  |
| --- | --- | --- | --- | --- | --- | --- |
|  | 2019 | 2020 | IRR<sup>a</sup> | 2019 | 2020 | IRR<sup>b</sup> |
| All procedures | 11,794 | 3,220 | 0.27 (0.25 to 0.3) | 19.28 | 12.39 | 0.64 (0.61 to 0.67) |
| Hysterectomy | 1,823 | 276 | 0.15 (0.13 to 0.18) | 57.95 | 25.58 | 0.4 (0.34 to 0.45) |
| Carotid endarterectomy | 476 | 121 | 0.26 (0.21 to 0.31) | 40.72 | 29.30 | 0.57 (0.48 to 0.66) |
| Coronary stenting | 5,675 | 1,646 | 0.29 (0.27 to 0.32) | 27.45 | 16.42 | 0.59 (0.56 to 0.63) |
| Spinal fusion or laminectomy | 1,649 | 322 | 0.2 (0.17 to 0.22) | 15.30 | 10.49 | 0.66 (0.59 to 0.73) |
| Vertebroplasty | 1,903 | 814 | 0.43 (0.39 to 0.47) | 8.57 | 7.34 | 0.85 (0.78 to 0.93) |
| Knee arthroscopy | 268 | 41 | 0.15 (0.1 to 0.23) | 8.01 | 10.17 | 0.89 (0.61 to 1.32) |
| All imaging/diagnostic services | 13,242 | 5,854 | 0.44 (0.43 to 0.46) | 9.09 | 8.45 | 0.93 (0.9 to 0.96) |
| EEG for syncope | 1,471 | 469 | 0.32 (0.29 to 0.36) | 2.18 | 1.38 | 0.63 (0.57 to 0.7) |
| Carotid imaging for syncope | 2,826 | 1,046 | 0.37 (0.34 to 0.4) | 6.52 | 4.87 | 0.74 (0.69 to 0.8) |
| EEG for headache | 182 | 73 | 0.4 (0.31 to 0.53) | 0.23 | 0.20 | 0.88 (0.67 to 1.16) |
| Head imaging for syncope | 10,378 | 4,864 | 0.47 (0.45 to 0.49) | 20.90 | 20.79 | 0.99 (0.96 to 1.03) |

<sup>a</sup>2019 vs 2020 incidence rate ratio (IRR) and 95% CI of patients with overuse services per Medicare beneficiaries; <sup>b</sup>2019 vs 2020 IRR and 95% CI of patients with overuse services per patients in overuse measure denominator cohort. EEG: Electroencephalography.

After April, the 2020 overuse procedure volumes were nearer to 2019 volumes (Figure 1). In July 2019 there were 11,525 patients with a low-value procedure versus 10,090 in 2020; IRR for rates per beneficiaries: 0.88 (95% CI 0.86 to 0.91; p <0.001). The overuse rates per measure denominators were also more like 2019: 19.00 patients per 100 beneficiaries in the July 2019 denominator cohort vs 18.79 in the July 2020 cohort, IRR 0.99 (95% CI 0.97 to 1.01; p = 0.339).

Several procedures had a decline in overuse rates in April 2020 compared to 2019 (Table 2), including coronary stenting, hysterectomy, carotid endarterectomy, and spinal fusion/laminectomy.

The number of patients with a low-value vertebroplasty also declined in April 2020 compared to 2019 (Table 2), and this was due to a drop in the number of patients included in the measure cohort (patients with an osteoporotic fracture) during this month: 11,088 patients in 2020 versus 22,217 patients in 2019. The overuse rate for these patients (that is, those who received a vertebroplasty) was only slightly lower than 2019 levels.

Knee arthroscopy overuse rates showed no significant change, likely due to the substantial drop in the overall volume of any knee arthroscopies (Table 2).

### Imaging/diagnostic volumes and overuse rates

There was a 55.58% decrease in the number of patients during April 2020 vs 2019 who received a low-value imaging/diagnostic service.

#### Syncope volumes and overuse rates

Three of the imaging overuse measures had the same denominator definition: patients with syncope (with some exclusions, see supplementary table 1). There were 49.45% fewer patients in April 2020 presenting with syncope than the previous year: 33,899 patients in 2020 versus 67,374 in 2019. By June 2020, the number of patients with syncope was 20.70% lower than 2019 levels.

The rates of these syncope patients receiving an EEG or carotid imaging service both declined during April 2020 (Table 2). Conversely, syncope patients received head imaging at the same rates in 2020 as 2019.

#### Headache volumes and overuse rates

The number of patients presenting to a facility with a headache also declined: there were 36,202 patients in April 2020 versus 80,027 in 2019. The overuse rate of these patients receiving an EEG did not change (Table 2).

The number of patients with a headache in 2020 were still lower than 2019 by December: 63,081 patients in April 2020 versus 83,057 in 2019, and there remained no difference in EEG overuse rates for this cohort (Figure 1).

### Service volumes and overuse rates by COVID-19 pandemic period

Figure 2 shows the 2020 versus 2019 IRRs for the service specific overuse rates by COVID-19 pandemic period. The largest decrease in volumes were during the initial shutdown period, ranging from an 81.11% decline in volumes of patients with low-value knee arthroscopy to a 52.85% decline in low-value head imaging for syncope patients.

**Figure 2.**
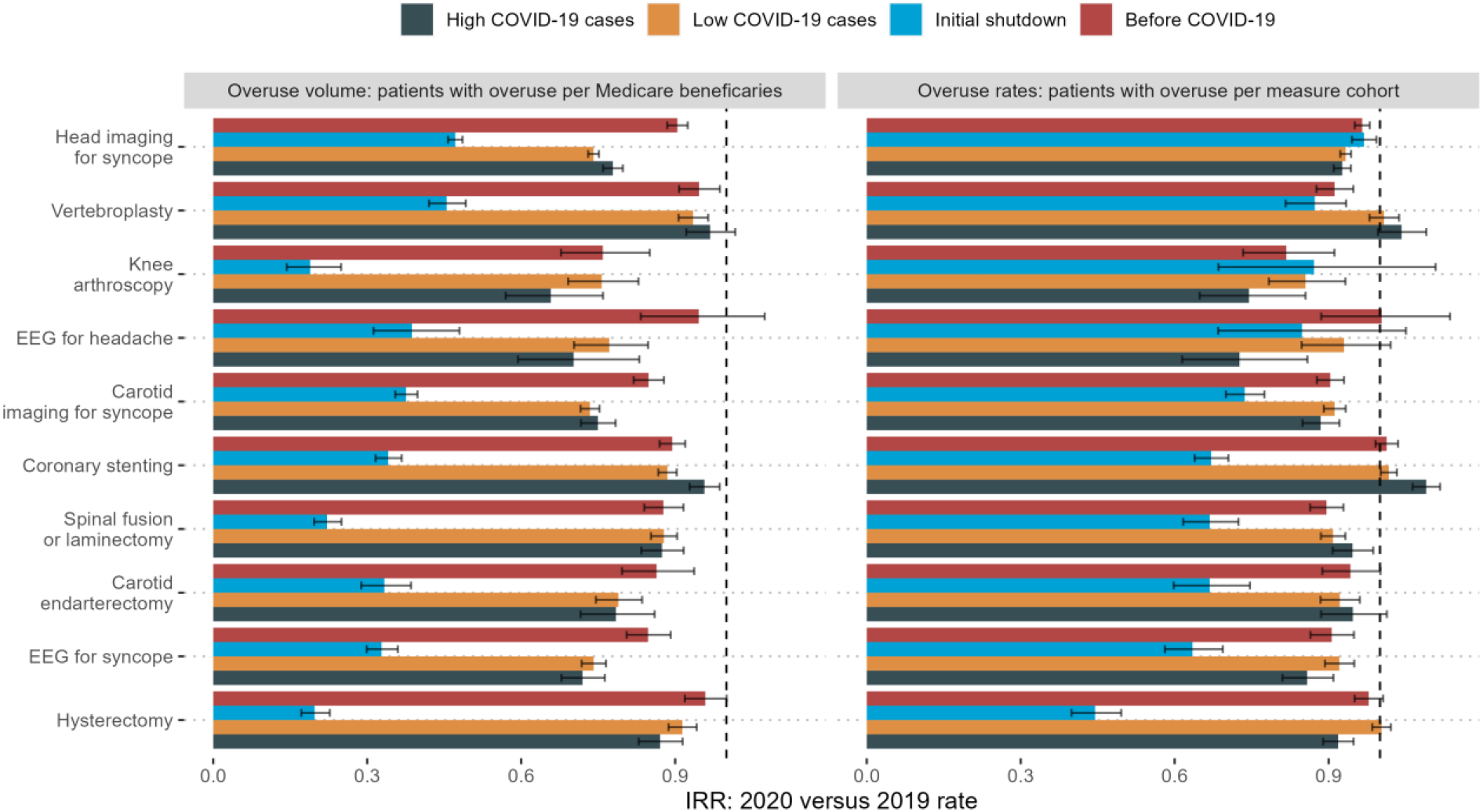
2020 versus 2019 incidence rate ratios for overuse measures by COVID-19 pandemic period. The incidence rate ratios (IRRs) for the number of patients with an overuse service per Medicare Fee-For-Service beneficiary (left panel) and per measure denominator cohort (right panel) in 2020 compared to 2019 in states prior to the pandemic (January, February), during the shutdown period (March 15 to May 2, 2020), periods of high COVID-19 incidence (greater than 175 cases per 100,000 persons) and periods of low COVID-19 incidence. *Measures where the cohort is defined as claims for patients with a certain condition are marked with an asterisk (vertebroplasty, EEG for syncope, carotid imaging for syncope, head imaging for syncope and EEG for headache patients). Other measures define the cohort as all patients with the service. IRRs and 95% CIs (error bars) were estimated from Poisson regressions by comparing total beneficiary counts during epidemiological weeks in 2020 with corresponding weeks in 2019.

Overuse rates also decreased for many services during the shutdown period, as Figure 1 previously showed. Hysterectomy had the largest change in proportions of overuse: 611 (28.21%) of 2,166 hysterectomies during the 2020 shutdown period were for patients with benign conditions, versus 3,118 (58.05%) of 5,371 hysterectomies during the same period in 2019 (IRR: 0.44 (95% CI 0.4 to 0.5; p <0.001)).

Outside the shutdown period, there were little differences in overuse rates between high COVID-19 incidence periods compared to low periods. In high COVID-19 periods, 54.09 per 100 hysterectomy patients were classed as overuse versus 57.48 per 100 hysterectomy patients in low COVID-19 periods, a decline of 8.15% and 0.29% respectively from the corresponding 2019 overuse rates (the comparative ratio of these rates was 0.92 (95% CI 0.88 to 0.95; p <0.001)). Similarly, EEG rates for patients with either syncope or headache was lower in high COVID-19 periods versus low COVID-19 periods; ratio for EEG for syncope was 0.93 (95% CI 0.87 to 1; p = 0.035) and for headache was 0.78 (95% CI 0.65 to 0.95; p = 0.011).

### Service volumes and overuse rates by COVID-19 pandemic period and state

Figure 3 shows the 2020 versus 2019 IRRs by the mean 7-day COVID-19 incidence for each state and COVID-19 incidence period. We omit head imaging for syncope as the Figure 1 results showed that this overuse measure rate did not change in 2020 relative to 2019.

**Figure 3.**
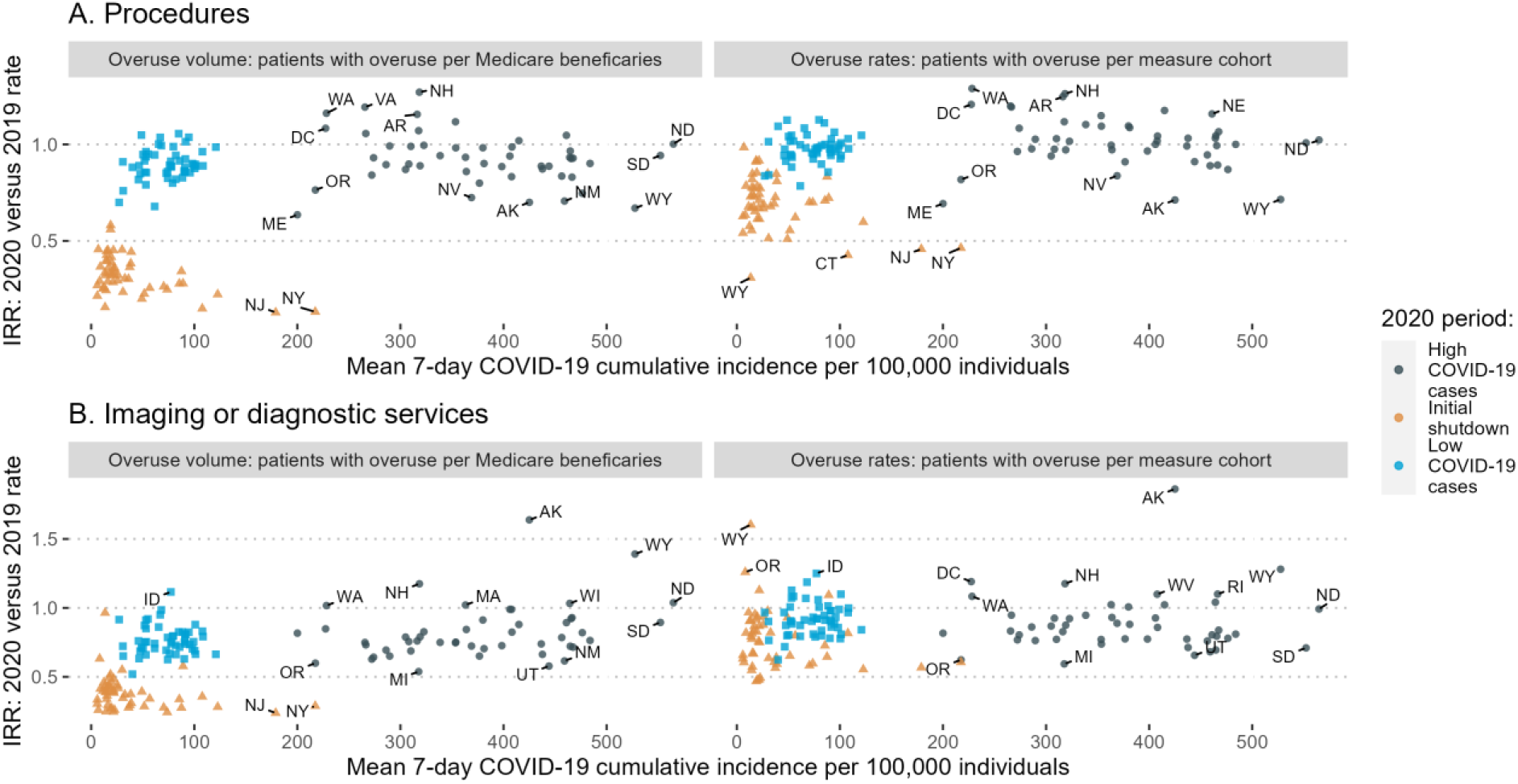
Stale 2020 versus 2019 incidence rate ratios for overuse measures by mean 7-day cumulative incidence of COVID-19 per population. Estimated incidence rate ratios (IRR) of rates in 2020 versus 2019 for each state versus the mean 7-day cumulative COVID-19 incidence rate. Left: IRRs for the number of patients with an overuse service per Medicare beneficiary; and Right: IRRs for the number of patients with an overuse service per measure denominator cohort. Panel A includes all procedure measures and panel B includes three imaging/diagnostic measures (EEG for headache, carotid imaging for syncope and EEG for syncope).

Procedure overuse rates and volumes had the greatest declines during the shutdown period compared to the rest of the 2020 pandemic period. New York, New Jersey, Massachusetts, and Connecticut had the highest mean 7-day cumulative COVID-19 incidence rates during this period and the largest decreases in number of overused procedures per beneficiary. The correlation between COVID-19 incidence and overuse volume IRRs was -0.52 (95% CI -0.70 to -0.29; p <0.001). The overuse rates also declined in states with mostly higher COVID-19 incidence rates (correlation -0.38 (95% CI -0.59 to -0.12; p = 0.006)). New Jersey, New York, Connecticut, and Wyoming had the largest decrease in procedure overuse rates during the shutdown period. Unlike the others, Wyoming had a low mean 7-day COVID-19 incidence during the shutdown period (13.44 cases per 100,000 population, opposed to New York’s 217.36 cases per 100,000 population).

After the shutdown period, changes in state procedure overuse rates relative to 2019 were not significantly correlated with COVID-19 incidence rates: correlation coefficients were -0.24 (95% CI -0.49 to 0.047; p = 0.101) for the high COVID-periods and 0.16 (95% CI -0.12 to 0.41; p = 0.274) for the low COVID periods.

The number of patients with an overused imaging/diagnostic service per beneficiary also declined during the shutdown period in each state. The state IRRs were negatively correlated with the mean COVID-19 incidences during the shutdown period (−0.28 (95% CI -0.51 to -0.0037; p = 0.047)). During high COVID-19 periods, however, there was actually a positive correlation between state overuse volume IRRs and COVID-19 incidence (0.31 (95% CI 0.029 to 0.54; p = 0.032)). There was no significant correlation between the 2020 vs 2019 IRRs for the imaging/diagnostic service overuse rates and the mean COVID-19 incidence in either low or high COVID-19 periods.

## Discussion

The disruption of business as usual at hospitals during the COVID-19 pandemic has served as a natural experiment providing a window on overuse practices.^9,10^ As expected based on previous research, the claim volume of procedures and certain conditions decreased during the initial COVID-19 pandemic shutdown. There was also a substantial decrease in overuse rates of most services during the shutdown period. Later in 2020, however, services overuse rates were mostly similar to previous rates prior to the pandemic and were not correlated with COVID-19 incidence.

Overuse is driven by multiple factors, some of which may be more discretionary than others.^11^ The changes in overuse rates observed during the shutdown period may be explained by government directives to delay non-urgent care.^12^ When the federal government declared a national emergency and hospitals were to provide urgent and necessary care only, fewer procedures and imaging/diagnostic services were provided. For example, we observed the proportion of coronary stents that were performed on patients with stable coronary disease declined during the shutdown period. This is presumably because these procedures were more likely to be delayed or canceled due to the shutdown compared to stenting for patients with myocardial infarction. The more urgent care will have taken priority.

Some services had a smaller or no decline in overuse rates during the shutdown period, including vertebroplasty for osteoporotic fractures, EEGs for headaches and head imaging for syncope. Although there was a decrease in the number of beneficiaries with claims for these conditions, the proportion of patients receiving an overuse procedure or imaging service remained like 2019 levels. There was no evidence that clinical decision making changed for these services.

There was evidence, however, that clinical decisions for syncope patients did change in the shutdown period. While these patients were just as likely to receive low-value head imaging as they were prior to the pandemic, they were less likely to receive a low value EEG or carotid imaging study. This might reflect the varying availability of imaging resources through the shutdown. Clinicians also may have delayed or decided against the use of EEG and carotid imaging studies, while head imaging (CT or MRIs) may have felt more compelling or accessible in this period.

The change in overuse rates of some services varied with high or low COVID-19 incidence. Hysterectomy and EEGs for headache or syncope all had greater declines in overuse rates during periods of high COVID-19 incidence compared to low COVID-19 incidence. Other services, such as spinal fusion/laminectomy, carotid endarterectomy, coronary stenting and vertebroplasty had no changes in overuse rates in high versus low COVID-19 incidence periods. This, and the fact that overuse volume IRR was positively associated with high COVID-19 incidence for imaging and diagnostic services, points to the possible discretionary use of some services during greater pandemic strain, but not other services.

Northeastern states had the most substantial state-specific decline in denominator cohorts and overuse rates during the shutdown period. These states all had the highest COVID-19 case incidence rates during this time, and the highest hospitalization and death rates.^13^ COVID-19 hospitalizations and/or death rates may predict impact on facility capacity (and therefore the denominator and overuse changes) more than case rates, particularly since in the early stages of the pandemic testing was less available than later in 2020 and cases may have been higher than reported.^14^

## Limitations

These overuse measures are indicators of low-value care and are limited by the diagnostic information available in claims data. We used measures from the Lown Institute overuse metric that had a specific denominator cohort.^7^ We did not include the two measures from this metric that have a denominator of all patients at the hospital over the selected period (inferior vena cava filters and renal stenting).

We adjusted for and investigated state-level beneficiary counts and COVID-19 incidence rates. A more granular approach could have used counts at the hospital referral region or county level and explored within and across region differences. COVID-19 incidence at more local regional levels may be more associated with overuse or patient volumes than at the state level. This could be future research built from this current study, which presents a more high-level overview of specific denominator and numerator overuse measure rates.

This study only investigated the impact on services through 2020. By the end of 2020, the US was entering another COVID-19 surge that lasted most of the winter. Later in 2021, the COVID-19 Omicron variant wave caused another surge in cases and hospitalizations. The changes to denominator and overuse rates of these measures may be different during each pandemic stage.

## Conclusions

For most investigated overuse measures, we observed the largest decrease in overuse rates during the COVID-19 shutdown period. These declines were larger than later in 2020 when COVID-19 case incidence was higher in the US. This suggests that the shutdown, an administrative intervention external to health care delivery, perhaps coupled with the initial uncertainty of the pandemic, had a larger impact on overuse than actual COVID-19 case levels. Notably, the overuse rates had all returned to 2019 levels shortly after the end of the shutdown period despite the ongoing pandemic. Seven out of the ten investigated measures had a significant decline in overuse rates as well as volume during the shutdown period, meaning that some selection or distinction of low-value care was apparent in clinical decision making for these measures compared to the others. The varying results, however, demonstrate that efforts to reduce low value care may require evaluations and interventions that are tailored to each service.

## Data Availability

Medicare data was accessed after approvals and agreement with CMS. Individual hospital results will be available on https://lownhospitalsindex.org/. Other data can be accessed upon reasonable request to The Lown Institute, please contact.

