## Supplement for "Overuse in US Medicare during the COVID-19 pandemic: 2020 versus 2019" (from another version of this preprint: version 1, 2022-05-26)

Supplementary information: overuse definitions

**Table** **1**: The overuse measure definitions for the claims included in the denominator and numerator (low-value) cohorts. Numerator criteria are applied in addition to the denominator criteria.

| **Service** | **Denominator** | **Numerator** |
| --- | --- | --- |
| Carotid endarterectomy (CEA) for female patients without a history of stroke or transient ischemic attack (TIA) and without stroke, TIA, or focal neurological symptoms noted in claim. Operation not associated with an emergency room (ER) visit.^1^ | CEA ICD-10-PCS codes in outpatient or MedPAR, or CEA CPT code in outpatient. In MedPAR, claim is included if this code is the principal procedure code. Exclude service if the beneficiary is male. | Exclude service if ICD-10-CM codes associated with stroke, TIA or focal neurological symptoms in claim. Exclude service if a patient’s first date of indication of Stroke/Transient Ischemic Attack from Chronic Conditions Warehouse (CCW) table is not empty and is prior to the service. Exclude service if a patient had any ER visits within 14 days prior. |
| Hysterectomy for benign disease^2^ | Hysterectomy ICD-10-PCS codes in outpatient or MedPAR, or CPT codes in outpatient. In MedPAR, claim is included if this code is the principal procedure code. | Exclude service if malignancy codes are in claim or carcinoma in situ codes. Exclude service if Diagnosis Related Group in MedPAR related to malignancy. |
| Arthroscopic debridement/ chondroplasty of the knee with diagnosis of osteoarthritis or chondromalacia in the procedure claim, and no meniscal tear^1^ | Arthroscopy ICD-10-PCS codes in outpatient or MedPAR, or CPT code in outpatient. In MedPAR, claim is included if this code is the principal procedure code. | Include service if osteoarthritis or chondromalacia is in claim. Exclude service if meniscus tear is in claim. |
| Coronary stent placement or balloon angioplasty for patients without unstable angina.^1^ Procedure not associated with an emergency room (ER) visit, which might be indicative of acute coronary syndrome. | ICD-10-PCS codes in outpatient or MedPAR, or CPT code in outpatient. In MedPAR, claim is included if this code is the principal procedure code. | Include service if the first indication date of Ischemic Heart Disease from the CCW data is at least 6 months prior to the service. Exclude service if any unstable angina or myocardial infarction codes in claim or within outpatient or MedPAR within the previous 14 days. Exclude service if a patient had any ER visits within 14 days prior. |
| Spinal fusion or laminectomy on lower vertebrae, excluding patients with radicular symptoms, trauma, herniated disc, discities, spondylosis, myelopathy, radiculopathy, radicular pain and scoliosis.^2–4^ | ICD-10-PCS codes in outpatient or MedPAR, or CPT code in outpatient. In MedPAR, claim is included if this code is the principal procedure code. | Exclude service if there are codes for cauda equina, congenital spondylolisthesis, disc displacement, discitis, hyperostosis, traumatic injury, lower limb pain, myelopathy, other disc disorders, paraplegia, postlaminectomy syndrome, radiculopathy, Schmorls nodes, sciatica, scoliosis, spondylosis, and other spine conditions Exclude spinal fusion cases if both lumbar stenosis with neurogenic claudication and lumbar spondylolisthesis is on the claim. |
| Vertebroplasty for vertebral fracture for patients with osteoporosis^1^ | ICD-10-PCS codes in outpatient or MedPAR, or CPT code in outpatient. In MedPAR, claim is included if this code is the principal procedure code or the principal procedure is kyphoplasty. | Include if osteoporosis pathological fracture code in MedPAR or Outpatient. Otherwise, include service if vertebra fracture code and patient has Osteoporosis indicator in CCW table prior to the service. Exclude claims with bone cancers, myeloma or hemangioma recorded. |
| EEG for headache, excluding patients with epilepsy or seizures^1^ | Include MedPAR and outpatient claims with headache. Exclude if both 1) headache is not the principal, admitting (MedPAR), or reason for admission diagnosis code (Outpatient) and 2) the beneficiary had headache diagnosis recorded in any claims in the prior two years. Exclude claims with epilepsy, recurrent seizures or abnormal involuntary movements. | Service is found in outpatient or MedPAR claims. |
| EEG for syncope^2^ | Include MedPAR and outpatient claims with syncope. Exclude if both 1) syncope is not the principal, admitting (MedPAR), or reason for admission diagnosis code (Outpatient) and 2) the beneficiary had syncope diagnosis recorded in any claims in the prior two years. | Service is found in outpatient or MedPAR claims. |
| Carotid imaging for syncope, with no stroke or stroke symptoms^1^ | Include MedPAR and outpatient claims with syncope. Exclude if both 1) syncope is not the principal, admitting (MedPAR), or reason for admission diagnosis code (Outpatient) and 2) the beneficiary had syncope diagnosis recorded in any claims in the prior two years. Exclude if the beneficiary has a prior Stroke / Transient Ischemic Attack flag present in CCW. Exclude if ICD-10-CM codes in claim relating to stroke, retinal vascular occlusions, nervous and musculoskeletal symptoms are present. | Service is found in outpatient or MedPAR claims. |
| Head imaging for syncope, excluding epilepsy, stroke or symptoms of stroke^1^ | Include MedPAR and outpatient claims with syncope. Exclude if both 1) syncope is not the principal, admitting (MedPAR), or reason for admission diagnosis code (Outpatient) and 2) the beneficiary had syncope diagnosis recorded in any claims in the prior two years. Exclude if ICD-10-CM codes in claim relating to epilepsy or convulsions, cerebrovascular diseases including stroke/TIA and subarachnoid hemorrhage, head or face trauma, altered mental status, nervous and musculoskeletal system symptoms, personal history of stroke/TIA are present. | Service is found in outpatient or MedPAR claims. |
| ICD-10-PCS: International Classification of Diseases, 10th edition, Procedure Coding System; ICD-10-CM: ICD-10 Clinical Modification; CPT: Current Procedural Terminology. | | |
